# A Randomized Phase 2/3 Study of Ensitrelvir, a Novel Oral SARS-CoV-2 3C-like Protease Inhibitor, in Japanese Patients With Mild-to-Moderate COVID-19 or Asymptomatic SARS-CoV-2 Infection: Results of the Phase 2a Part

**DOI:** 10.1101/2022.05.17.22275027

**Authors:** Hiroshi Mukae, Hiroshi Yotsuyanagi, Norio Ohmagari, Yohei Doi, Takumi Imamura, Takuhiro Sonoyama, Takahiro Fukuhara, Genki Ichihashi, Takao Sanaki, Keiko Baba, Yosuke Takeda, Yuko Tsuge, Takeki Uehara

## Abstract

For the treatment of coronavirus disease 2019 (COVID-19), antiviral agents that can achieve rapid severe acute respiratory syndrome coronavirus 2 (SARS-CoV-2) reduction are warranted. This double-blind, phase 2a part of a phase 2/3 study assessed the efficacy and safety of ensitrelvir, a novel oral SARS-CoV-2 3C-like protease inhibitor, in Japanese patients with mild-to-moderate COVID-19 or asymptomatic SARS-CoV-2 infection. Sixty-nine patients enrolled from 56 sites were randomized (1:1:1) to orally receive 5-day ensitrelvir fumaric acid (375 mg on day 1 followed by 125 mg daily or 750 mg on day 1 followed by 250 mg daily) or placebo and followed up until day 28. The primary outcome was change from baseline in SARS-CoV-2 viral titer. A total of 16, 14, and 17 patients in the ensitrelvir 125 mg, ensitrelvir 250 mg, and placebo groups, respectively, were included in the intention-to-treat population (mean age: 38.8, 40.4, and 38.0 years, respectively). On day 4, the change from baseline in SARS-CoV-2 viral titer (log_10_ 50% tissue culture infectious dose/mL) in patients with positive viral titer and viral RNA at baseline was greater with ensitrelvir 125 mg (mean [standard deviation], −2.42 [1.42]; *P* = 0.0712) and 250 mg (−2.81 [1.21]; *P* = 0.0083) versus placebo (−1.54 [0.74]), and ensitrelvir treatment reduced SARS-CoV-2 RNA by −1.4 to −1.5 log_10_ copies/mL versus placebo. All adverse events were mild to moderate. Ensitrelvir treatment demonstrated rapid SARS-CoV-2 clearance and was well tolerated in patients with mild-to-moderate COVID-19 or asymptomatic SARS-CoV-2 infection (Japan Registry of Clinical Trials identifier: jRCT2031210350).

## INTRODUCTION

The coronavirus disease 2019 (COVID-19) outbreak, caused by infection with severe acute respiratory syndrome coronavirus 2 (SARS-CoV-2), has quickly spread worldwide and poses a significant public health burden. As of April 11, 2022, nearly 500 million confirmed cases of COVID-19 and more than 6 million deaths associated with COVID-19 have been reported to the World Health Organization (https://covid19.who.int/). Although at least one-third of SARS-CoV-2 infections are asymptomatic (1), the risk of severe COVID-19 is high in approximately one-fifth of infected individuals, particularly in those with underlying health conditions (2). Shedding of infectious SARS-CoV-2 may persist for approximately 10 days in asymptomatic individuals or patients with mild-to-moderate COVID-19 and up to 4 weeks in those with severe disease after symptom onset (3–5). A high viral titer or long persistence of infectious SARS-CoV-2 affects not only the disease severity of infected patients but may also facilitate its spread to their close contacts. Moreover, although vaccination is the cornerstone for controlling COVID-19, it is occasionally associated with post-vaccination breakthrough infections (6). Therefore, from a public health point of view, rapid SARS-CoV-2 reduction in infected patients, in addition to vaccination, is essential for disease control.

Several novel treatment options have demonstrated efficacy in reducing the risk of hospitalization, death, or disease progression in patients with COVID-19 at risk of severe disease. These include molnupiravir, a small-molecule ribonucleoside prodrug of *N*-hydroxycytidine (7); nirmatrelvir, a SARS-CoV-2 3C-like (3CL) protease inhibitor, in combination with ritonavir as a pharmacokinetic booster (8); the SARS-CoV-2 neutralizing monoclonal antibody sotrovimab (9); and the neutralizing antibody cocktail casirivimab/imdevimab (10). In addition, remdesivir has demonstrated efficacy in patients hospitalized with COVID-19, with an acceptable safety profile (11). Additional oral antiviral agents that can achieve rapid SARS-CoV-2 reduction as home-based treatment are required for patients with mild-to-moderate COVID-19 or asymptomatic SARS-CoV-2 infection.

Ensitrelvir fumaric acid (S-217622; hereafter, ensitrelvir) is a novel oral SARS-CoV-2 3CL protease inhibitor that was discovered through joint research by Hokkaido University and Shionogi & Co., Ltd. (12) and is currently developed by Shionogi & Co., Ltd. for the treatment of COVID-19. As the SARS-CoV-2 3CL protease is essential for processing viral complex proteins needed for viral replication (13), its inhibition by ensitrelvir is expected to prevent viral replication. Indeed, ensitrelvir has shown antiviral efficacy in both *in vitro* and *in vivo* animal studies (12, 14). Moreover, its safety, tolerability, and pharmacokinetic profile as single and multiple oral doses have been assessed in a phase 1 study (R. Shimizu, et al., unpublished data). Currently, a multicenter, randomized, double-blind, placebo-controlled, phase 2/3 study is underway to assess the efficacy, safety, and pharmacokinetics of 5-day oral administration of ensitrelvir (Japan Registry of Clinical Trials identifier: jRCT2031210350). Herein, we report the results of the phase 2a part of this phase 2/3 study that assessed the antiviral efficacy and safety of ensitrelvir in patients with mild-to-moderate COVID-19 or asymptomatic SARS-CoV-2 infection.

## RESULTS

### Patient disposition

This phase 2a part was conducted from September 28, 2021, to January 1, 2022, at 56 sites across Japan (Table S1). Of the 70 patients who provided informed consent and were screened, 1 was excluded prior to randomization due to protocol deviation. Among the 69 patients randomized (ensitrelvir 125 mg, 22; ensitrelvir 250 mg, 23; and placebo, 24), 1 patient in the ensitrelvir 125 mg group did not receive the study drug and was excluded from the safety analysis set. After receiving the allocated intervention, 3 patients in the placebo group discontinued treatment because of consent withdrawal (2 patients) or disease progression (1 patient). None of the female patients discontinued treatment because of pregnancy. Twenty-two patients who tested negative with reverse transcription-polymerase chain reaction (RT-PCR) at baseline (ensitrelvir 125 mg, 6; ensitrelvir 250 mg, 9; and placebo, 7) were excluded, resulting in 47 patients (ensitrelvir 125 mg, 16; ensitrelvir 250 mg, 14; and placebo, 17) in the intention-to-treat (ITT) population. Following the exclusion of patients without detectable baseline viral titer, 43 patients (ensitrelvir 125 mg, 15; ensitrelvir 250 mg, 14; and placebo, 14) were included in the modified ITT (mITT) population (Fig. 1). All analyses were performed in originally assigned groups.

**FIG 1.**
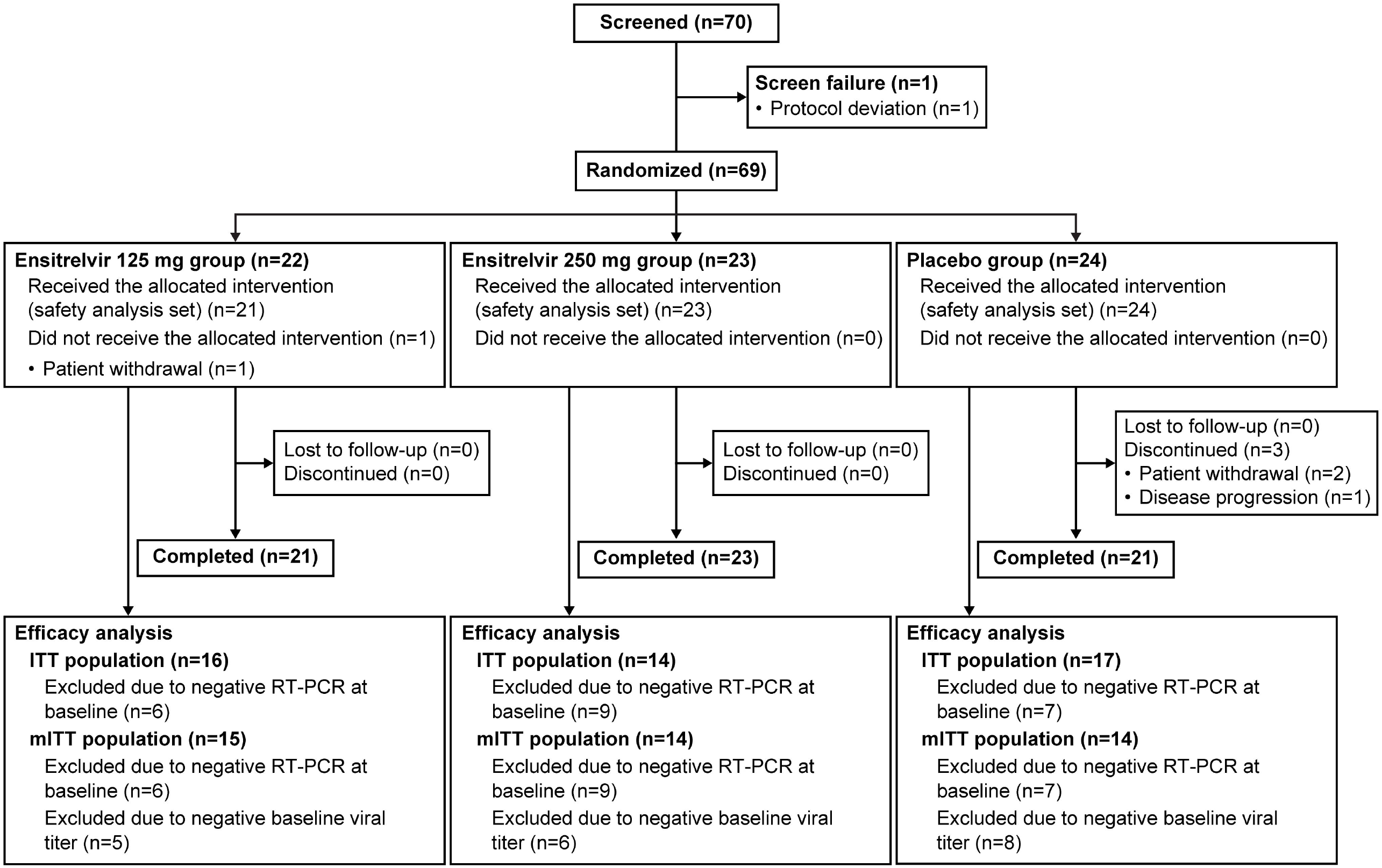
Patient disposition Patients may be excluded from the analysis populations due to more than one reason. ITT, intention-to-treat; mITT, modified ITT; RT-PCR, reverse transcription-polymerase chain reaction.

### Demographics and clinical characteristics

No notable difference was observed in the baseline demographics and clinical characteristics across treatment groups (Table 1). The mean (standard deviation [SD]) age among the 47 patients included in the ITT population was 38.8 (12.5), 40.4 (10.7), and 38.0 (14.2) years in the ensitrelvir 125 mg, ensitrelvir 250 mg, and placebo groups, respectively. None of the enrolled patients were aged > 65 years, and those aged < 20 years were enrolled in the placebo group only. Of the 47 patients, 29 (ensitrelvir 125 mg, 8 [50.0%]; ensitrelvir 250 mg, 8 [57.1%]; and placebo, 13 [76.5%]) were men. A majority of the ITT population (40 patients) had mild-to-moderate COVID-19, and 7 (ensitrelvir 125 mg, 2 [12.5%]; ensitrelvir 250 mg, 2 [14.3%]; and placebo, 3 [17.6%]) were asymptomatic. Among the 47 patients, 42 and 5 were infected with the Delta and Omicron variants, respectively. A majority of the patients in the ITT population had been vaccinated (ensitrelvir 125 mg, 14 [87.5%]; ensitrelvir 250 mg, 12 [85.7%]; and placebo, 12 [70.6%]). All vaccinated patients had received 2 doses of mRNA SARS-CoV-2 vaccines, except for 1 patient in the ensitrelvir 125 mg group who received only 1 dose. All patients in the ITT population completed the 5-day treatment, except for 1 patient each in the ensitrelvir 125 mg group (did not receive the study drug) and placebo group (discontinued the treatment after day 2).

**TABLE 1.**
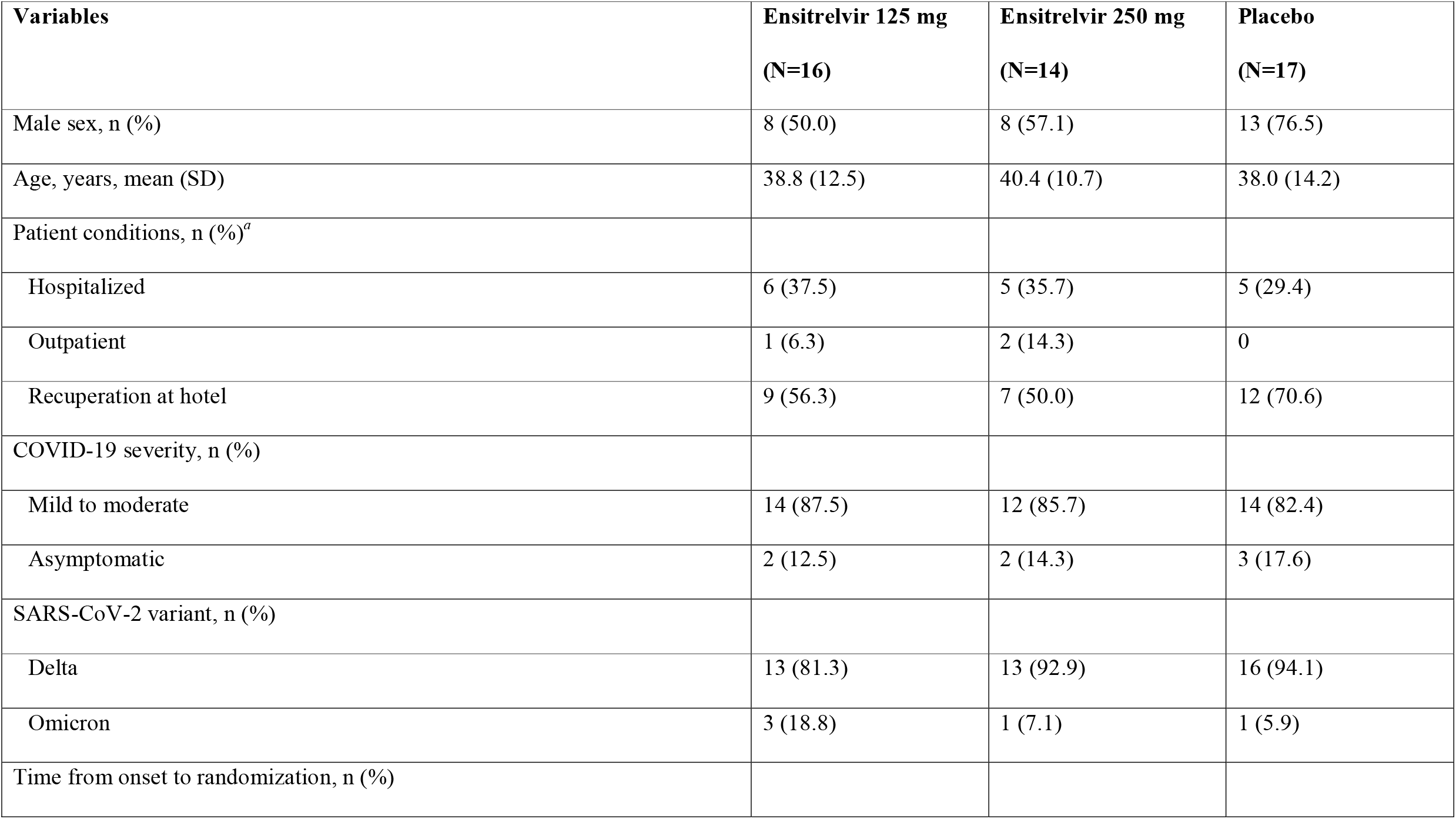
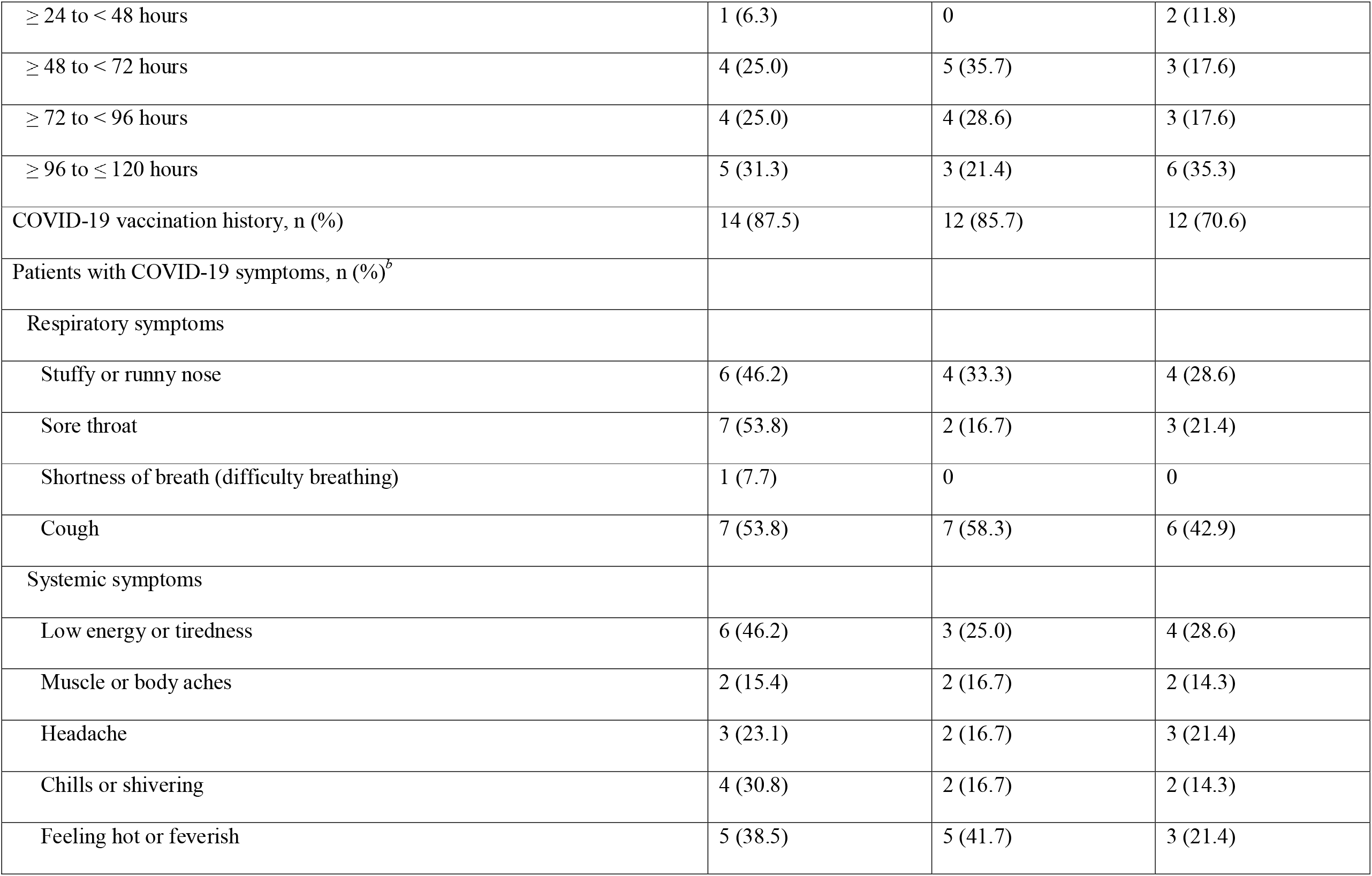
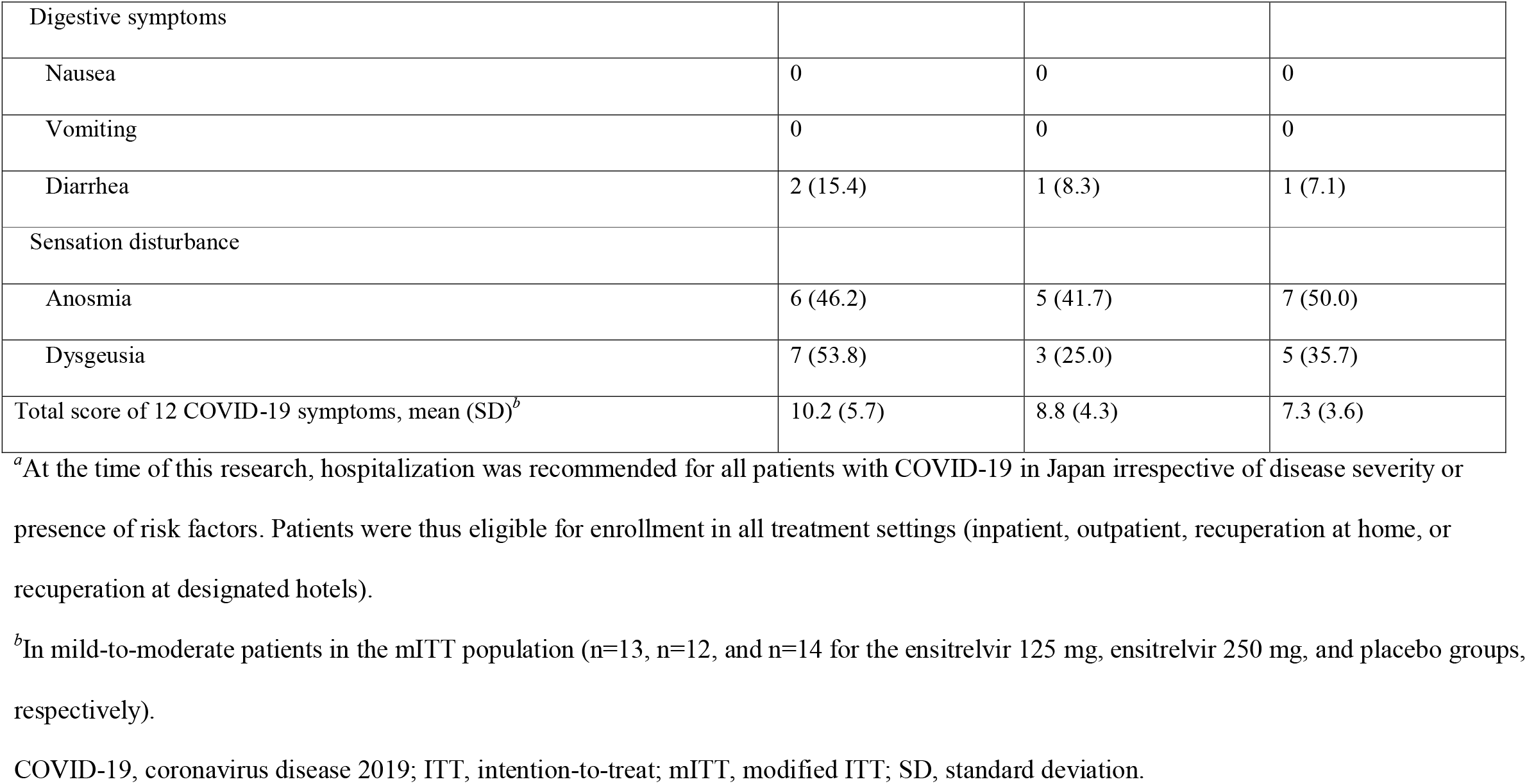
Baseline demographics and clinical characteristics (ITT population)

### SARS-CoV-2 viral titer and viral RNA level

Generally, the SARS-CoV-2 viral titer, measured as log_10_ 50% tissue culture infectious dose (TCID_50_)/mL, decreased with time for up to day 4 (3 days after treatment initiation) and remained stable until day 21 in all treatment groups. The SARS-CoV-2 viral titer was significantly lower in the ensitrelvir 125 mg (mean [SD], 0.94 [0.29]; *P* = 0.0333) and 250 mg (0.85 [0.19]; *P* = 0.0059) groups versus the placebo group (1.74 [1.17]) on day 4 (Fig. 2A). The change from baseline in SARS-CoV-2 viral titer on day 4 was greater in the ensitrelvir 125 mg (mean [SD], −2.42 [1.42]; difference from placebo, −0.88; *P* = 0.0712) and 250 mg (−2.81 [1.21]; difference from placebo, −1.27; *P* = 0.0083) groups versus the placebo group (−1.54 [0.74]) (Fig. 2B).

**FIG 2.**
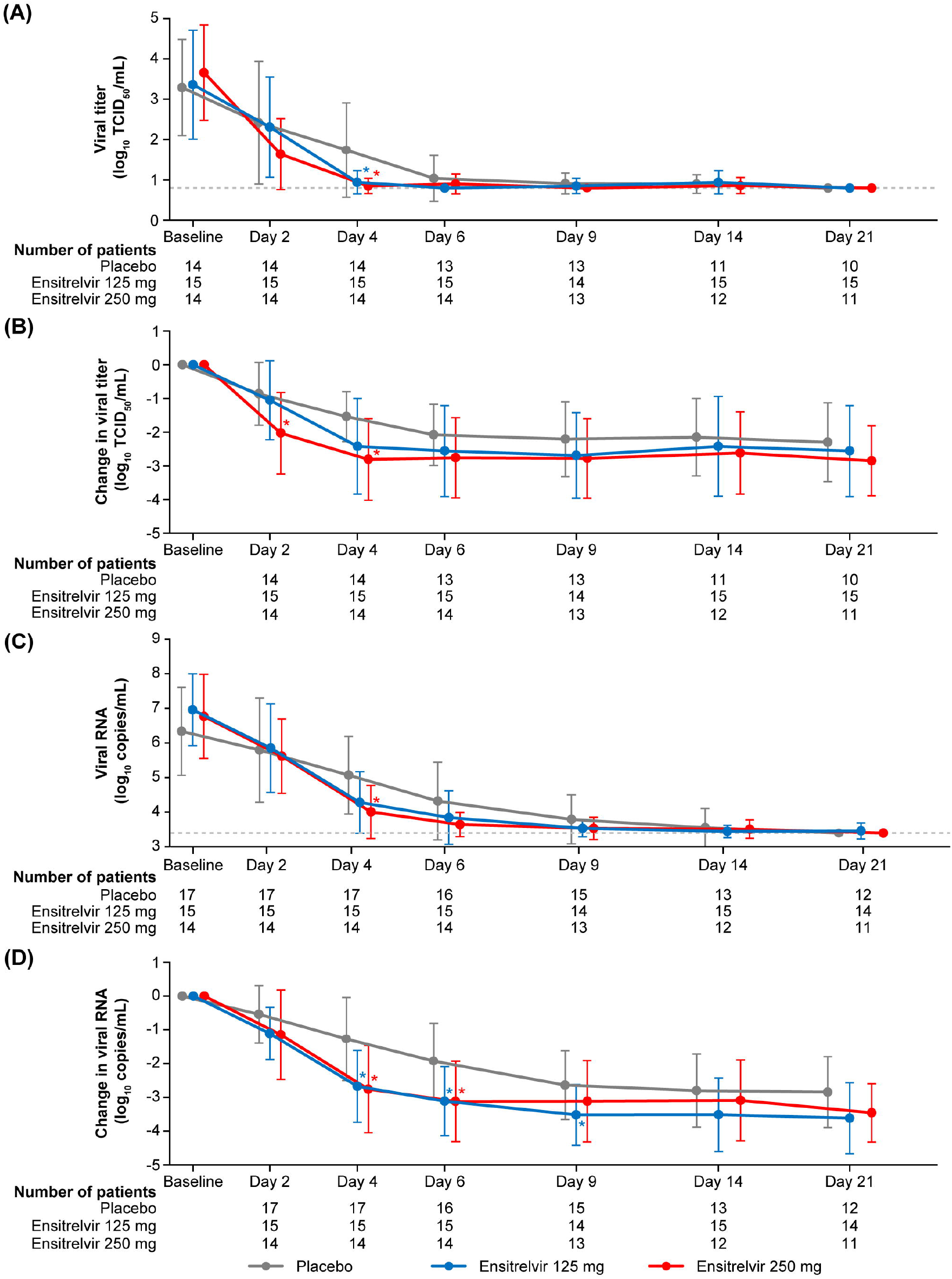
SARS-CoV-2 viral titer as (A) absolute values and (B) change from baseline (mITT population), and SARS-CoV-2 viral RNA level as (C) absolute values and (D) change from baseline (ITT population) Data are presented as mean ± SD. \**P* < 0.05 versus placebo. The gray, dotted, horizontal lines in panels (A) and (C) indicate the lower limit of detection of viral titer (0.8 log_10_ TCID_50_/mL) and lower limit of quantification of viral RNA (3.40 log_10_ copies/mL), respectively. ITT, intention-to-treat; mITT, modified ITT; SARS-CoV-2, severe acute respiratory syndrome coronavirus 2; SD, standard deviation; TCID_50_, 50% tissue culture infectious dose.

The SARS-CoV-2 viral RNA level, measured using RT-PCR (log_10_ copies/mL), was significantly lower in the ensitrelvir 250 mg group versus the placebo group on day 4 (mean [SD], 4.005 [0.765] versus 5.067 [1.117]; *P* = 0.0084) (Fig. 2C). The change from baseline in viral RNA level (log_10_ copies/mL) on day 4 was significantly greater in the ensitrelvir 125 mg (mean [SD], −2.677 [1.063]; difference from placebo, −1.408; *P* = 0.0029) and 250 mg (−2.761 [1.291]; difference from placebo, −1.492; *P* = 0.0039) groups versus the placebo group (−1.269 [1.228]). A similar trend of greater change in viral RNA level versus the placebo group was observed on day 6 in the ensitrelvir 125 mg (*P* = 0.0108) and 250 mg (*P* = 0.0041) groups and day 9 in the ensitrelvir 125 mg group (*P* = 0.0323) (Fig. 2D).

### Proportion of patients with positive SARS-CoV-2 viral titer

The proportion of patients with positive SARS-CoV-2 viral titer decreased with time from baseline to day 21 in all treatment groups. On day 4, the proportion of patients with positive viral titer was significantly lower in the ensitrelvir 125 mg (26.7%, *P* = 0.0205) and 250 mg (14.3%, *P* = 0.0033) groups versus the placebo group (71.4%), with a reduction of approximately 60% and 80%, respectively. Similarly, the proportion of patients with positive viral titer was significantly lower in the ensitrelvir 125 mg group versus the placebo group (0% versus 30.8%, *P* = 0.0276) on day 6 (Fig. S1).

### Time to viral clearance

The median time to viral clearance (first negative SARS-CoV-2 viral titer) was significantly shorter in the ensitrelvir 125 mg (61.3 hours; difference from placebo, −49.8 hours; *P* = 0.0159) and 250 mg (62.7 hours; difference from placebo, −48.4 hours; *P* = 0.0205) groups versus the placebo group (111.1 hours). Viral clearance was achieved in all patients in the ensitrelvir 125 mg and 250 mg groups in less than 120 hours (Fig. 3).

**FIG 3.**
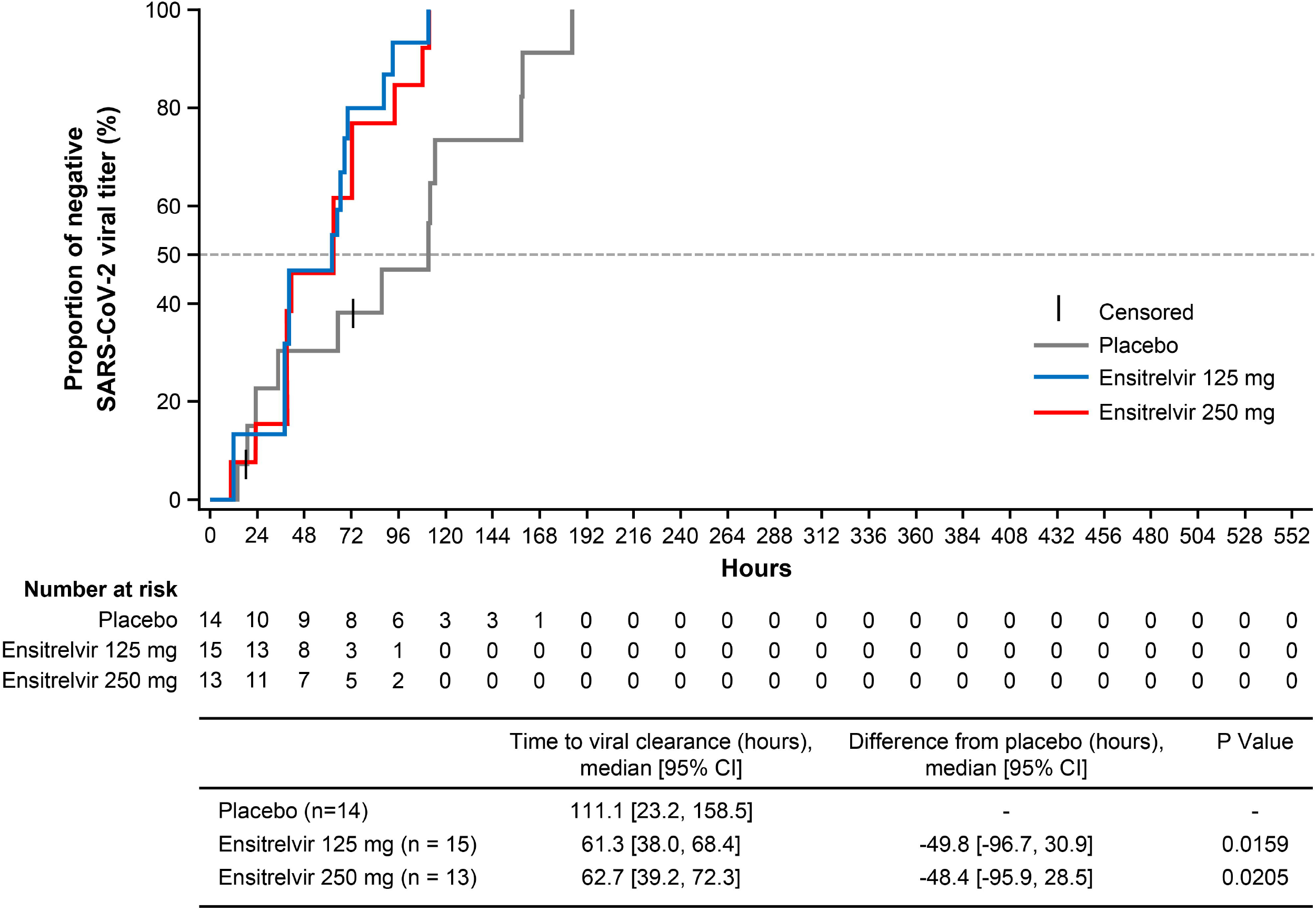
Time to viral clearance (first negative SARS-CoV-2 viral titer; mITT population) One patient in the ensitrelvir 250 mg group was excluded from this analysis due to the use of prohibited concomitant drugs on day 1. CI, confidence interval; mITT, modified intention-to-treat; SARS-CoV-2, severe acute respiratory syndrome coronavirus 2.

### COVID-19 symptoms

The mean total score of the predefined 12 symptoms in patients with mild-to-moderate COVID-19 showed a decreasing trend with time after treatment initiation in all groups (Fig. 4A). Generally, the mean change from baseline in the total score of the 12 COVID-19 symptoms was numerically greater in the ensitrelvir 125 mg and 250 mg groups versus the placebo group (Fig. 4B). A decreasing trend from baseline to 120 hours after dose in the 14 COVID-19 symptom scores was observed. In particular, the scores for respiratory symptoms (stuffy or runny nose, sore throat, and cough) and feverish symptoms showed a decreasing trend in patients treated with ensitrelvir (Fig. S2).

**FIG 4.**
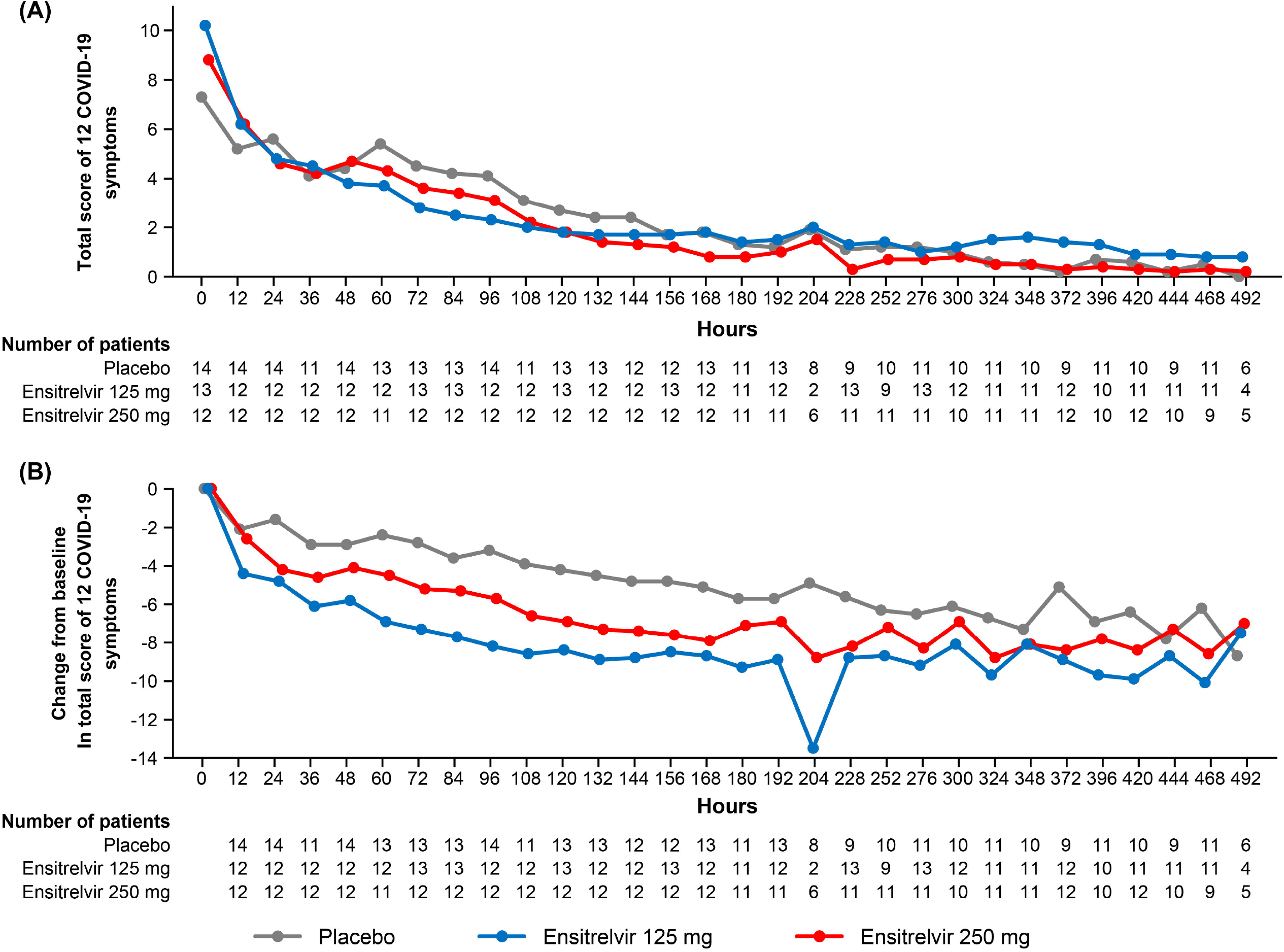
Total score of 12 COVID-19 symptoms in patients with mild-to-moderate COVID-19 as (A) mean absolute value and (B) mean change from baseline (ITT population) COVID-19, coronavirus disease 2019; ITT, intention-to-treat.

### Effects in preventing COVID-19 exacerbations (*post hoc* analysis)

After treatment initiation, 2 of 14 patients in the placebo group recorded ≥3 on the 8-point ordinal scale of patients’ condition (exacerbation), both of whom were unvaccinated. None of the patients in the ensitrelvir 125 mg (unvaccinated, 2; vaccinated, 11) and 250 mg (unvaccinated, 2; vaccinated, 10) groups recorded ≥3 on the 8-point ordinal scale (Table S2).

### Safety

Treatment-emergent adverse events (TEAEs) were reported in 11 (52.4%), 16 (69.6%), and 9 (37.5%) patients in the ensitrelvir 125 mg, ensitrelvir 250 mg, and placebo groups, respectively, all of which were mild to moderate in severity. The most common TEAE observed across groups was a decrease in high density lipoprotein (HDL; 3 [14.3%], 12 [52.2%], and 2 [8.3%] patients in the ensitrelvir 125 mg, ensitrelvir 250 mg, and placebo groups, respectively) (Table 2). Transient decrease and increase in HDL and blood triglyceride levels, respectively, were observed in the ensitrelvir groups following treatment initiation but were resolved without additional treatment (Fig. S3). Additionally, an increase in total bilirubin and iron levels was observed in patients receiving ensitrelvir (Fig. S4). Total bilirubin increase observed in 2 patients in the ensitrelvir 250 mg group (Table 2) and iron increase observed in 1 patient in the ensitrelvir 250 mg group were reported as TEAEs. These changes were asymptomatic and transient, and resolved without additional treatment. No obvious changes compared with placebo were observed in haptoglobin, reticulocytes, and low-density lipoprotein in the ensitrelvir groups, and no laboratory or clinical signs of hemolysis were observed. No serious TEAEs, deaths, or TEAEs leading to treatment discontinuation were reported (Table 2).

**TABLE 2.** Summary of TEAEs (safety analysis set)

| **Patients with events, n (%)** | **Ensitrelvir 125 mg**<br>**(N=21)** | **Ensitrelvir 250 mg**<br>**(N=23)** | **Placebo**<br>**(N=24)** |
| --- | --- | --- | --- |
| Patients with any TEAE | 11 (52.4) | 16 (69.6) | 9 (37.5) |
| Patients with any serious TEAE or death | 0 (0.0) | 0 (0.0) | 0 (0.0) |
| Patients with TEAEs leading to treatment discontinuation | 0 (0.0) | 0 (0.0) | 0 (0.0) |
| TEAEs occurring in $\geq 5\%$ of patients in either group | | | |
| Nasopharyngitis | 2 (9.5) | 0 (0.0) | 0 (0.0) |
| Headache | 1 (4.8) | 3 (13.0) | 0 (0.0) |
| Rhinalgia | 2 (9.5) | 0 (0.0) | 0 (0.0) |
| High density lipoprotein decreased | 3 (14.3) | 12 (52.2) | 2 (8.3) |
| Blood triglycerides increased | 0 (0.0) | 3 (13.0) | 0 (0.0) |
| Aspartate aminotransferase increased | 1 (4.8) | 1 (4.3) | 2 (8.3) |
| Blood bilirubin increased | 0 (0.0) | 2 (8.7) | 0 (0.0) |
| Alanine aminotransferase increased | 1 (4.8) | 0 (0.0) | 2 (8.3) |
| Patients with any treatment-related AE | 5 (23.8) | 10 (43.5) | 0 (0.0) |
| Treatment-related AEs occurring in $\geq 5\%$ of patients in either group | | | |
| High density lipoprotein decreased | 3 (14.3) | 8 (34.8) | 0 (0.0) |
| Blood triglycerides increased | 0 (0.0) | 2 (8.7) | 0 (0.0) |

10 AE, adverse event; TEAE, treatment-emergent AE

Treatment-related AEs were reported in 5 (23.8%) and 10 (43.5%) patients in the ensitrelvir 125 mg and 250 mg groups, respectively. A decrease in HDL and increase in blood triglycerides were observed to be the most common (≥5% in either group) treatment-related AEs (Table 2). All treatment-related AEs resolved without sequelae, except for mild hyperbilirubinemia (total bilirubin level just above the normal range) that lasted until the last visit (day 28) in 1 patient in the 250 mg group.

## DISCUSSION

This is the first clinical study to assess the antiviral efficacy and safety of a novel oral SARS-CoV-2 3CL protease inhibitor, ensitrelvir, in patients with mild-to-moderate COVID-19 or asymptomatic SARS-CoV-2 infection. This study used infectious viral titer as the primary endpoint to assess the antiviral efficacy of anti-SARS-CoV-2 agents. A majority of the enrolled patients had been vaccinated, which reflects the real-world scenario. Our results demonstrated promising antiviral efficacy of 5-day oral administration of ensitrelvir in rapid viral titer and viral RNA reduction in this patient population. Patients with mild-to-moderate COVID-19 who were treated with ensitrelvir showed a trend of symptom relief. Treatment with ensitrelvir was generally well tolerated.

As patients with COVID-19 may shed infectious viruses for up to 4 weeks depending on the disease severity (3–5), rapid viral clearance is crucial for disease treatment. In the current phase 2a part, reduction in viral titer and viral RNA was observed after ensitrelvir treatment; this efficacy is comparable with or greater than the findings from clinical trials for other anti-SARS-CoV-2 infection agents (7, 8, 15). Moreover, viral RNA reduction with ensitrelvir was substantial, with a difference from placebo on day 4 reported at −1.4 to −1.5 log_10_ copies/mL. The difference from placebo in viral RNA reduction reported in previous randomized clinical trials on day 5 was −0.547 log_10_ copies/mL (least-squares mean) for molnupiravir 800 mg (15) and −0.868 log_10_ copies/mL (adjusted mean) for nirmatrelvir plus ritonavir (8). In addition, previous randomized clinical trials of anti-SARS-CoV-2 agents were conducted in unvaccinated patients (7, 8, 15), whereas most patients in our study had been vaccinated. These results indicate the efficacy of ensitrelvir in the rapid clearance of SARS-CoV-2. From the efficacy of ensitrelvir in rapid viral titer and viral RNA reduction, favorable clinical outcomes, such as earlier symptom relief or prevention of severe COVID-19, are expected.

Transient decrease in HDL and increase in triglycerides was observed as TEAEs in the current phase 2a part, which was consistent with the findings derived from the previous phase 1 study of ensitrelvir (R. Shimizu, et al., unpublished data). In addition, transient and asymptomatic increase in total bilirubin and iron levels, which was not associated with signs of hepatic or hematologic abnormalities, was observed following ensitrelvir treatment. However, the exact mechanism of these laboratory value changes is yet to be elucidated and further investigation is warranted.

This study has some limitations. First, the small sample size and high vaccination rate limit concrete conclusions to be drawn from analyses of the clinical outcomes, such as COVID-19 symptoms including respiratory symptoms and disease exacerbation. Second, most of the enrolled patients were infected with the SARS-CoV-2 Delta variant. Our *in vitro* study results suggested that ensitrelvir has antiviral activity against the Omicron variant, which is the most predominant SARS-CoV-2 variant at the time of publication (12). Thus, the clinical efficacy of ensitrelvir against the Omicron variant or future variants of concern needs to be assessed in the subsequent part of this study. In addition, statistical analyses in subgroups, such as minor patients and asymptomatic patients, were not feasible owing to the limited number of patients. Further assessment of the safety and clinical efficacy of ensitrelvir in subsequent clinical studies is warranted.

In conclusion, treatment with 5-day oral administration of ensitrelvir demonstrated a rapid clearance of SARS-CoV-2 and was well tolerated in patients with mild-to-moderate COVID-19 or asymptomatic SARS-CoV-2 infection. The results support further clinical development of ensitrelvir through large-scale clinical studies for the treatment of mild-to-moderate COVID-19 or asymptomatic SARS-CoV-2 infection.

## MATERIALS AND METHODS

### Study design

Patients with mild-to-moderate COVID-19 or asymptomatic SARS-CoV-2 infection were randomized (1:1:1) to receive ensitrelvir fumaric acid tablet 125 mg, 250 mg, or matching placebo. Patients allocated to the ensitrelvir groups received a loading dose of ensitrelvir on day 1 (375 mg for the 125 mg group and 750 mg for the 250 mg group), followed by the maintenance dose (125 mg for the 125 mg group and 250 mg for the 250 mg group) on days 2 through 5 without dose modification.

This study was conducted in accordance with the principles of the Declaration of Helsinki, Good Clinical Practice guidelines, and other applicable laws and regulations. The study was reviewed and approved by the institutional review boards of all participating institutions listed in Table S1. All patients or their legally acceptable representatives provided written informed consent.

### Patients

Patients (aged 12 to <70 years; body weight ≥40 kg for those aged <20 years to avoid increased drug exposure) who tested positive for SARS-CoV-2 within 120 hours prior to randomization were eligible for study enrollment. Results of SARS-CoV-2 antigen tests or nucleic acid detection testing, including RT-PCR, were used to determine positive SARS-CoV-2 infection. Patients with mild-to-moderate COVID-19 should have had at least 1 moderate or severe symptom among the 12 COVID-19 symptoms (stuffy or runny nose, sore throat, shortness of breath, cough, low energy or tiredness, muscle or body aches, headache, chills or shivering, feeling hot or feverish, nausea, vomiting, or diarrhea; Table S3), based on the United States Food and Drug Administration guidance for assessing COVID-19-related symptoms (https://www.fda.gov/regulatory-information/search-fda-guidance-documents/assessing-covid-19-related-symptoms-outpatient-adult-and-adolescent-subjects-clinical-trials-drugs), or worsening of at least 1 moderate or severe COVID-19 symptom. Patients without the 12 COVID-19 symptoms listed above, anosmia, or dysgeusia within 2 weeks prior to randomization were classified as those with asymptomatic SARS-CoV-2 infection.

Key exclusion criteria were an awake oxygen saturation of ≤93% (room air); need for oxygen administration; worsening of COVID-19 within 48 hours from randomization strongly suspected by the investigator; suspected active and systemic infections requiring treatment (except for COVID-19); current or chronic history of moderate or severe liver disease, known hepatic or biliary abnormalities (except for Gilbert syndrome or asymptomatic gallstones), or kidney disease; recent blood donation (≥400 mL within 12 weeks or ≥200 mL within 4 weeks prior to enrollment); use of drugs for COVID-19 within 7 days; and use of strong CYP3A inhibitors or inducers or St John’s wort products within 14 days prior to randomization. Pregnant, possibly pregnant, or breastfeeding women were also excluded.

### Randomization and blinding

Randomization of patients was performed using an interactive response technology system. For patients with mild-to-moderate COVID-19, the time from the onset of COVID-19 to randomization (<72 hours/≥72 hours) and the presence or absence of COVID-19 vaccination were used as stratification factors. For those with asymptomatic SARS-CoV-2 infection, the presence or absence of COVID-19 vaccination was used as a stratification factor.

All patients and study staff were blinded to treatment until the completion of the 28-day follow-up period and database lock, except for designated persons at the sponsor and contract research organization in charge of statistical analyses. Measurements of SARS-CoV-2 viral titer and viral RNA level prior to database lock were allowed in an anonymized manner to avoid unexpected unblinding. Emergency unblinding per the investigator’s request was allowed only in the event of AEs to determine an appropriate therapy for the patient.

### Treatment

Patients received allocated study drugs orally (ensitrelvir fumaric acid 125 mg, 250 mg, or placebo tablets), which were indistinguishable in appearance, labeling, and packaging.

Use of drugs for the treatment of COVID-19; antiviral, antibacterial, or antifungal drugs (except for external use); antipyretic analgesics other than acetaminophen; antitussives and expectorants; combination cold remedy; and CYP3A substrates was prohibited from study initiation to day 28 or study discontinuation. Strong CYP3A inhibitors or inducers, strong P-glycoprotein or breast cancer resistance protein inhibitors, and other transporter substrates were prohibited from study initiation to 10 days after the last administration of the study drug or study discontinuation. Treatment was discontinued in the event of exacerbation of COVID-19, serious or intolerable AEs, pregnancy, or liver function abnormalities.

### Outcomes and assessments

The primary outcome was a change from baseline (day 1, before drug administration) in SARS-CoV-2 viral titer on days 2, 4, 6, 9, 14, and 21 (or study discontinuation). Secondary outcomes included SARS-CoV-2 viral RNA level, proportion of patients with a positive viral titer, and time to viral clearance (first negative SARS-CoV-2 viral titer). SARS-CoV-2 viral titer measurement and RT-PCR using nasopharyngeal swabs were performed at Shionogi TechnoAdvance Research (Osaka, Japan) and LSI Medience (Tokyo, Japan), respectively.

Clinical outcomes included the total score of the 12 COVID-19 symptoms, proportion of patients with anosmia, and those with dysgeusia following treatment in patients with mild-to-moderate COVID-19. Patients assessed their COVID-19 symptoms on a questionnaire (Table S3) and recorded the scores in a diary twice daily (morning and evening) until day 9 and once daily (evening) from days 10 to 21. Additionally, disease exacerbations were assessed as a *post hoc* analysis in patients with mild-to-moderate COVID-19. Patients’ conditions (exacerbation) were assessed by the investigator using an 8-point ordinal scale (0 = Asymptomatic to 7 = Death, ≥3 corresponds to hospitalization or worse symptoms; Table S4) on day 1 (before drug administration) and days 2, 4, 6, 9, 14, 21, and 28 (or study discontinuation).

Safety was assessed through TEAEs coded using Medical Dictionary for Regulatory Activities version 24.0. Laboratory tests, vital sign measurements, and electrocardiography were additionally performed. All safety data were evaluated by an independent data and safety monitoring board. Pregnancy tests were performed on day 1 (before drug administration), day 28, and at the investigator’s discretion for women of childbearing potential.

### Statistical analyses

To compare change from baseline in SARS-CoV-2 viral titer between each treatment group and the placebo group, 11 patients per group with COVID-19 were required to provide 82.3% power, assuming a difference of 2.5 log, a common SD of 2.2 log, and a 2-sided significance level of 10%. Based on the phase 2a study of molnupiravir, which reported an infectious SARS-CoV-2 rate of 44% at baseline (15), it was assumed that 50% of patients enrolled in this phase 2a part would have positive RT-PCR results and detectable viral titer at baseline. Thus, the sample size was set to 23 patients per group (69 in total).

All randomized patients who tested positive with RT-PCR at baseline were included in the ITT population. All randomized patients who tested positive with RT-PCR and had detectable SARS-CoV-2 viral titer at baseline were included in the mITT population. All randomized patients who received at least 1 dose of the study drug were included in the safety analysis set.

A positive SARS-CoV-2 viral titer was defined as ≥0.8 log_10_ TCID_50_/mL. SARS-CoV-2 viral titer and SARS-CoV-2 viral RNA level were compared between each of the ensitrelvir groups and placebo using the van Elteren test. Median time to viral clearance was compared between each of the ensitrelvir groups and placebo using a stratified log-rank test. The proportion of patients with positive SARS-CoV-2 viral titer was compared between each of the ensitrelvir groups and placebo using the Mantel-Haenszel test. All statistical tests were performed with stratification by study cohort (mild to moderate or asymptomatic) and at a 2-sided significance level of 0.05.

No prespecified statistical hypothesis was set because this phase 2a part was conducted in an exploratory manner. No imputation was performed for missing data, and multiplicity was not considered. All analyses were performed using SAS version 9.4 (SAS Institute, Inc., Cary, NC, USA).

## Data Availability

Shionogi & Co., Ltd. is committed to disclosing the synopses and results of its clinical trials and sharing clinical trial data with researchers on reasonable request. For further details, please refer to the websites of Shionogi & Co., Ltd. (https://www.shionogi.com/shionogi/global/en/company/policies/shionogi-group-clinical-trial-data-transparency-policy.html) and Vivli (https://vivli.org/).

## ACKNOWLEDGMENTS

The authors and research team thank all patients involved in this study and Masahiro Kinoshita and Satoshi Kojima (Shionogi & Co., Ltd.) for their support for manuscript development. Support for study monitoring and data management was provided by EPS Corporation and funded by Shionogi & Co., Ltd. Medical writing and editorial assistance was provided by Mami Hirano, MS, of Cactus Life Sciences (part of Cactus Communications) and funded by Shionogi & Co., Ltd. All authors retained full ownership of the manuscript content and approved the final draft for submission.

This study was sponsored by Shionogi & Co., Ltd. and financially supported by the Japan Agency for Medical Research and Development. Employees of Shionogi & Co., Ltd. participated in and approved the design and conduct of the study, wrote the protocol, and were involved in the collection, management, analysis, and interpretation of data. Institutional authors reviewed and approved the protocol and collected and interpreted the data.

H. Mukae has received funding relevant to the submitted work from Shionogi and grants from Taisho Pharma; lecture fees from Pfizer, MSD, Shionogi, and Taisho Pharma; and advisory fees from Pfizer, MSD, and Shionogi outside the submitted work. H. Yotsuyanagi has received consulting fees from Shionogi, lecture fees from Shionogi and ViiV Healthcare, and travel support from Shionogi outside the submitted work. He serves as an advisory board member of Shionogi and President of the Japanese Society of Infectious Diseases. Y. Doi has received grants from Shionogi and Entasis; consulting fees from Shionogi, Meiji Seika Pharma, Gilead Sciences, GSK, MSD, Chugai, and bioMerieux; and lecture fees from MSD, AstraZeneca, Shionogi, and Teijin Healthcare outside the submitted work and serves as an advisory board member of FujiFilm. T. Imamura, T. Sonoyama, T. Fukuhara, G. Ichihashi, T. Sanaki, K. Baba, Y. Takeda, Y. Tsuge, and T. Uehara are full-time employees of Shionogi & Co., Ltd. and may have stocks or stock options. N. Ohmagari declares no conflict of interest.

## Author contributions

H. Mukae: Conceptualization, Data curation, Supervision, Writing – review & editing. H. Yotsuyanagi: Conceptualization, Data curation, Writing – review & editing. N. Ohmagari: Conceptualization, Data curation, Writing – review & editing. Y. Doi: Conceptualization, Data curation, Writing – review & editing. T. Imamura: Conceptualization, Formal analysis, Supervision, Visualization, Writing – review & editing. T. Sonoyama: Conceptualization, Supervision, Writing – review & editing. T. Fukuhara: Conceptualization, Data curation, Project administration, Writing – review & editing. G. Ichihashi: Conceptualization, Data curation, Project administration, Writing – review & editing. T. Sanaki: Conceptualization, Data curation, Formal analysis, Methodology, Writing – review & editing. K. Baba: Conceptualization, Data curation, Formal analysis, Methodology, Writing – review & editing. Y. Takeda: Conceptualization, Data curation, Project administration, Writing – review & editing. Y. Tsuge: Conceptualization, Project administration, Supervision, Writing – review & editing. T. Uehara: Conceptualization, Project administration, Supervision, Writing – review & editing.

