## Supplementary material for "A Randomized Phase 2/3 Study of Ensitrelvir, a Novel Oral SARS-CoV-2 3C-like Protease Inhibitor, in Japanese Patients With Mild-to-Moderate COVID-19 or Asymptomatic SARS-CoV-2 Infection: Results of the Phase 2a Part": FIG S1 to S4, TABLE S1 to S4

**Supplemental material**

**FIG S1** Proportion of patients with positive SARS-CoV-2 viral titer (mITT population)

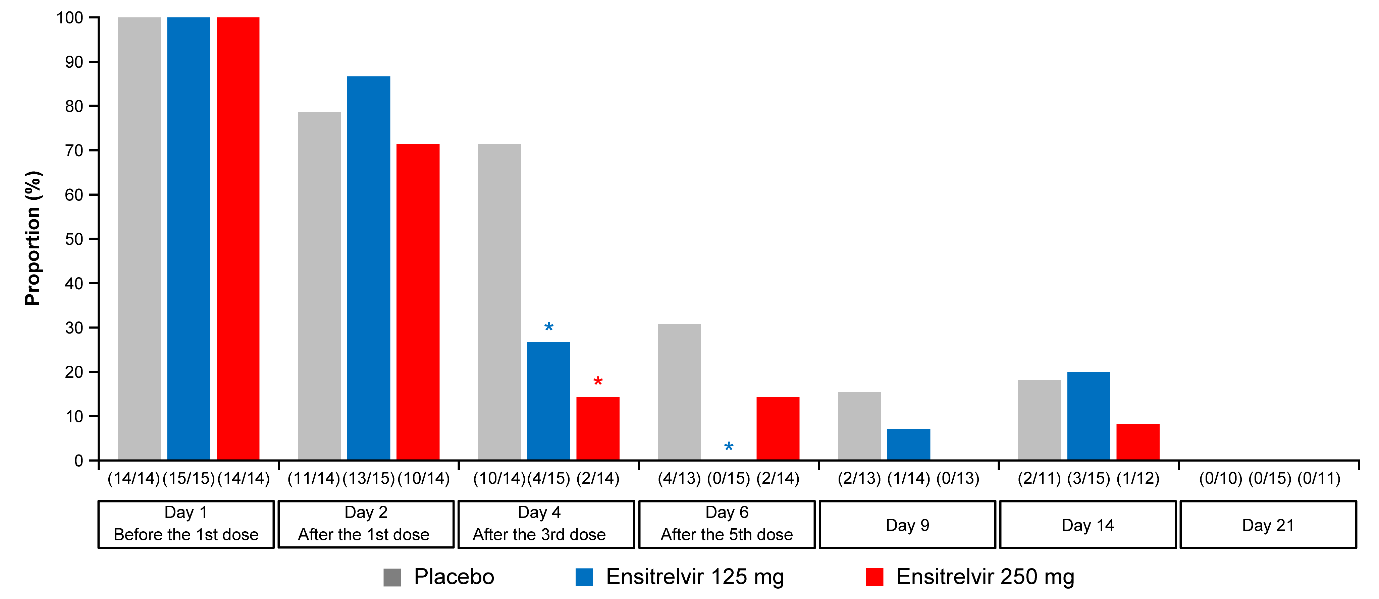

**P* < 0.05 versus placebo.

mITT, modified intention-to-treat; SARS-CoV-2, severe acute respiratory syndrome coronavirus 2.

**FIG S2** Mean change from baseline in each of the 14 COVID-19 symptom scores (ITT population)

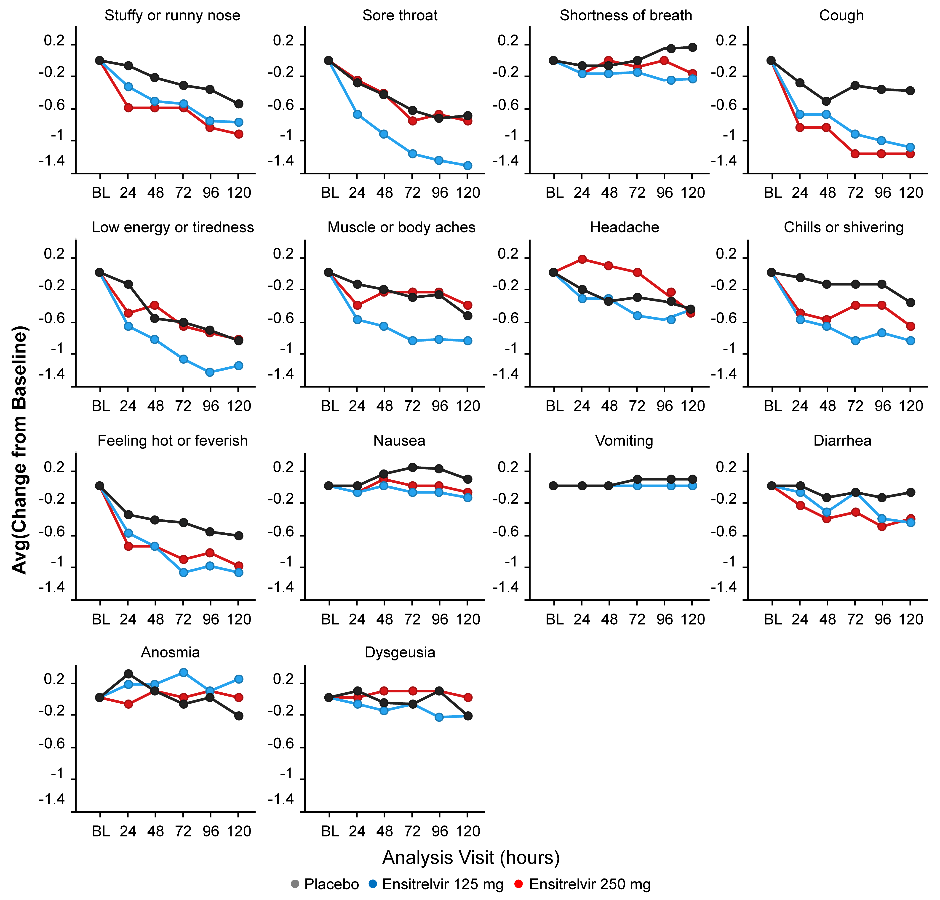

BL, baseline; COVID-19, coronavirus disease 2019; ITT, intention-to-treat.

**FIG S3** HDL cholesterol and blood triglyceride levels as (A) absolute values and (B) change from baseline (safety analysis set)

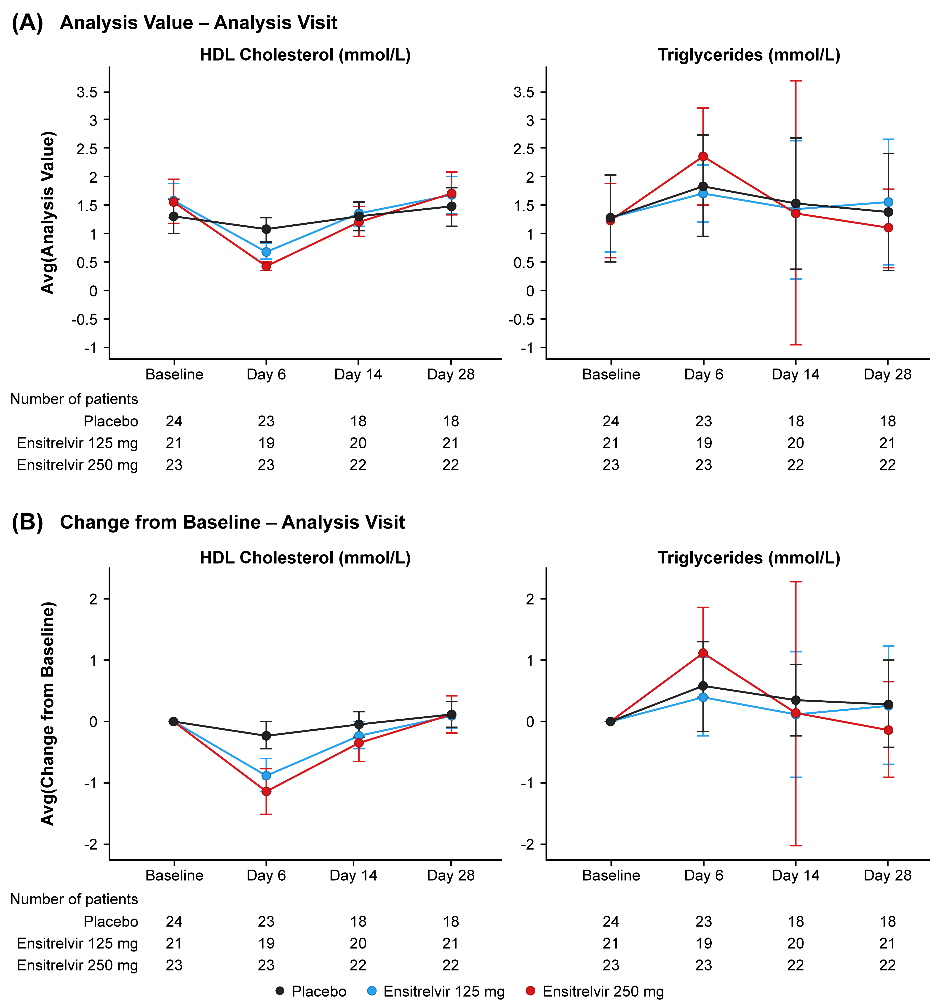

Data are presented as mean ± SD.

HDL, high density lipoprotein; SD, standard deviation.

**FIG S4 Total bilirubin and iron levels** as (A) absolute values and (B) change from baseline (safety analysis set)

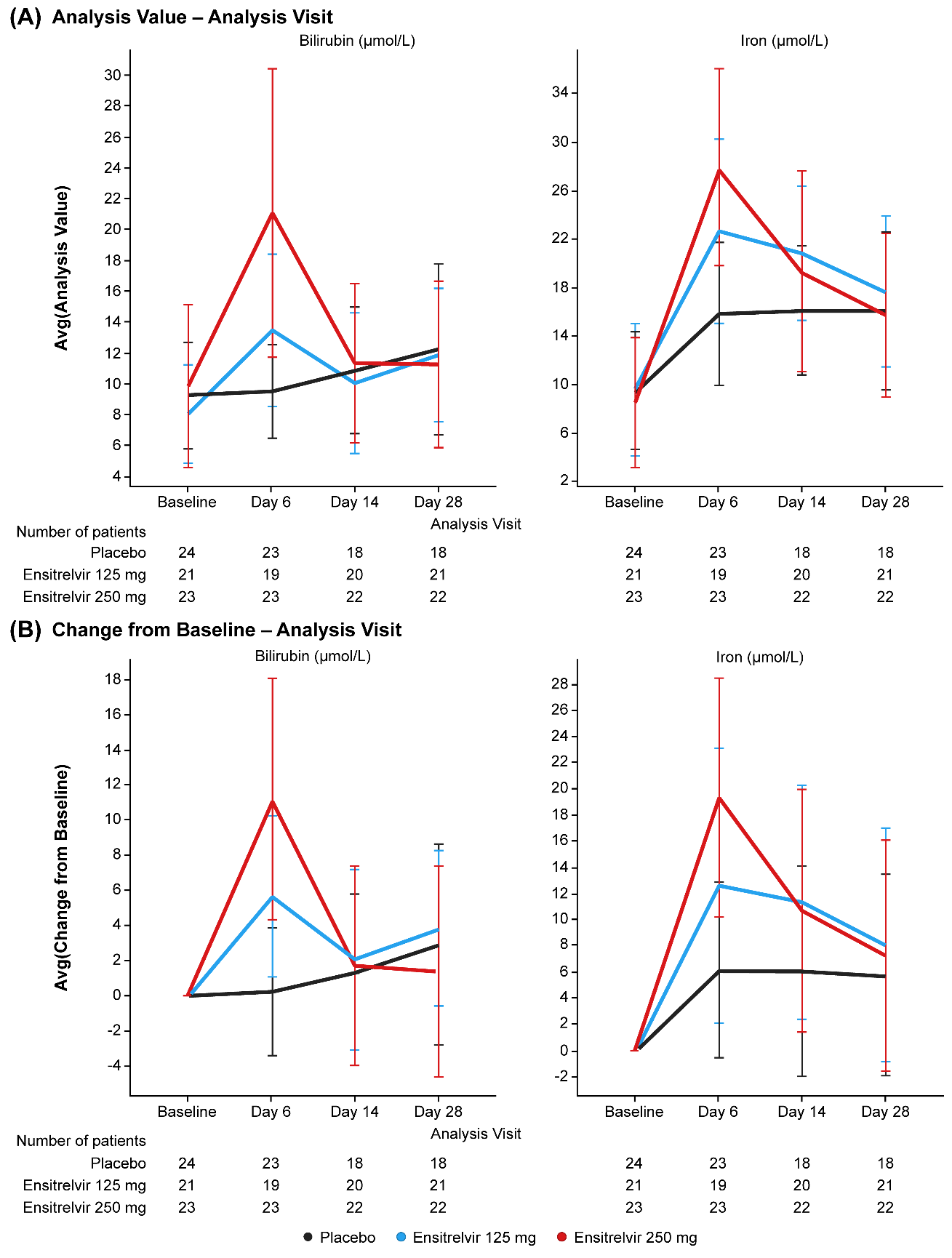

Data are presented as mean ± SD.

SD, standard deviation.

**TABLE S1** List of participating institutions and institutional review boards

| **Institution** | **Institutional review board** |
| --- | --- |
| **Japan** | |
| Kodaira Hospital | Sugiura Clinic Institutional Review Board |
| Swing Nozaki Clinic |  |
| Kasukabe Medical Center |  |
| Japan Community Health care Organization Tokyo Shinjuku Medical Center |  |
| Kouwakai Kouwa Clinic |  |
| Onga Nakama Medical Association Onga Hospital |  |
| Tsuchiura Beryl Clinic |  |
| Edogawa Medicare Hospital |  |
| Kamezawa Clinic |  |
| Ginza Sawada Clinic | Adachi Kyosai Hospital Institutional Review Board |
| Yaguchi Midori Clinic |  |
| Kanagawa Himawari Clinic |  |
| Minna no Tennocho Clinic |  |
| Matsunami General Hospital |  |
| Japan Community Health care Organization Shimonoseki Medical Center |  |
| Irie Clinic |  |
| Fukuoka Shinmizumaki Hospital |  |
| Tashiro Thyroid Clinic |  |
| JA Toride Medical Center | Review Board of Human Rights and Ethics for Clinical Studies Institutional Review Board |
| Moriya Keiyu Hospital |  |
| Japan Community Health care Organization Osaka Minato Central Hospital |  |
| Kinki Central Hospital of the Mutual Aid Association of Public School Teachers |  |
| Medical Corporation Kouhoukai Takagi Hospital |  |
| Fujimaki Ent Clinic | Yoga Allergy Clinic Institutional Review Board |
| Kaiseikai Kita Shin Yokohama Internal Medicine Clinic |  |
| Lee Clinic |  |
| Koga General Hospital | Tokushukai Group Institutional Review Board |
| Kamagaya General Hospital |  |
| Nozaki Tokushukai Hospital |  |
| Ikoma City Hospital |  |
| Showa University Hospital | Showa University Hospital Institutional Review Board |
| Showa University East Hospital |  |
| Yokohama Municipal Citizen’s Hospital | Yokohama Municipal Citizen’s Hospital Institutional Review Board |
| Aso Iizuka Hospital | Aso Iizuka Hospital Institutional Review Board |
| Red Cross Kyoto Daini Hospital | Japanese Red Cross Society Japanese Red Cross Kyoto Daini Hospital Institutional Review Board |
| Tokyo Metropolitan Health and Hospitals Corporation Toshima Hospital | Tokyo Metropolitan Health and Hospitals Toshima Hospital Institutional Review Board |
| IUHW Narita Hospital | International University of Health and Welfare and Institutional Review Board |
| Center Hospital of the National Center for Global Health and Medicine | Center Hospital of National Center for Global Health and Medicine IRB |
| Denenchofu Family Clinic | Kobori Central Clinical Research Ethics Committee |
| Osaka Rosai Hospital | Osaka Rosai Hospital Institutional Review Board |
| Kojunkai Daido Medicine | Daido Hospital Institutional Review Board |
| Rinku General Medical Center | Rinku General Medical Center IRB |
| University of Tsukuba Hospital | Institutional Review Board, University of Tsukuba Hospital |
| Nagasaki University Hospital | Nagasaki University Hospital Institutional Review Board |
| Tokyo Medical University Hachioji Medical Center | Tokyo Medical University Hachioji Medical Center Institutional Review Board |
| IMSUT Hospital, The Institute of Medical Science, The University of Tokyo | IMSUT Hospital, The Institute of Medical Science, The University of Tokyo Institutional Review Board |
| Fujita Health University Okazaki Medical Center | The Central Institutional Review Board for the Fujita Health University Hospitals |
| National Hospital Organization Yokohama Medical Center | National Hospital Organization Yokohama Medical Center Institutional Review Board |
| Okinawa National Hospital | Okinawa National Hospital Clinical Trial Review Committee |
| National Hospital Organization Kobe Medical Center | National Hospital Organization Kobe Medical Center Institutional Review Board |
| National Hospital Organization Himeji Medical Center | National Hospital Organization Himeji Medical Center Institutional Review Board |
| National Hospital Organization Fukuoka National Hospital | National Hospital Organization Fukuoka National Hospital Institutional Review Board |
| National Hospital Organization Chiba Medical Center | National Hospital Organization Chiba Medical Center Institutional Review Board |
| Japan Community Health Care Organization Hokkaido Hospital | Review Board of Japan Community Health care Organization Hokkaido Hospital |
| Hamamatsu Medical Center | Hamamatsu Medical Center Institutional Review Board |
| Nagoya City University East Medical Center | NCU East/West Medical Center Institutional Review Board |

**TABLE S2** Proportion of patients with mild-to-moderate COVID-19 who had disease exacerbation (≥3 in the 8-point ordinal scale) after treatment initiation (ITT population)

| **Patients** | **Ensitrelvir 125 mg**  **(N=13)** | **Ensitrelvir 250 mg**  **(N=12)** | **Placebo**  **(N=14)** |
| --- | --- | --- | --- |
| Patients with exacerbation, n/N (%) |  |  |  |
| Overall | 0/13 (0.0) | 0/12 (0.0) | 2/14 (14.3) |
| Unvaccinated | 0/2 (0.0) | 0/2 (0.0) | 2/4 (50.0) |
| Vaccinated | 0/11 (0.0) | 0/10 (0.0) | 0/10 (0.0) |

Data are derived from mild-to-moderate patients in the ITT population.

COVID-19, coronavirus disease 2019; ITT, intention-to-treat.

**TABLE S3** Questionnaire for the COVID-19 symptom scores

| **Questionnaire item** | **Response options and scoring** |
| --- | --- |
| Respiratory symptoms | None=0  Mild=1  Moderate=2  Severe=3 |
| 1. Stuffy or runny nose*^a^* |  |
| 2. Sore throat*^a^* |  |
| 3. Shortness of breath (difficulty breathing)*^a^* |  |
| 4. Cough*^a^* |  |
| Systemic symptoms |  |
| 5. Low energy or tiredness*^a^* |  |
| 6. Muscle or body aches*^a^* |  |
| 7. Headache*^a^* |  |
| 8. Chills or shivering*^a^* |  |
| 9. Feeling hot or feverish*^a^* |  |
| Digestive symptoms |  |
| 10. Nausea (feeling like you wanted to throw up)*^a^* |  |
| 11. Vomiting (throwing up) |  |
| 12. Diarrhea (loose or watery stools) |  |
| Sensation disturbance | Same as usual=0  Less than usual=1  No sense of smell/taste=2 |
| 13. Rate your sense of smell in the last 24 hours*^b^* |  |
| 14. Rate your sense of taste in the last 24 hours*^b^* |  |

*^a^*Patients were asked to rate the severity of their symptoms at their worst over the last 24 hours.

*^b^*Not used to calculate the total score of the 12 COVID-19 symptoms.

COVID-19, coronavirus disease 2019.

**TABLE S4** The 8-point ordinal scale for patients’ conditions

| **Descriptor** | **Score** |
| --- | --- |
| Asymptomatic | 0 |
| Symptomatic, no limitation of activities | 1 |
| Symptomatic, limitation of activities | 2 |
| Hospitalized, no oxygen therapy | 3 |
| Hospitalized, with oxygen therapy (<5 L/min) | 4 |
| Hospitalized, with oxygen therapy (≥5 L/min) | 5 |
| Hospitalized, with ventilation | 6 |
| Death | 7 |
